# DNA methylation mediates the effect of cocaine use on HIV severity

**DOI:** 10.1101/2020.05.11.20027458

**Authors:** Chang Shu, Amy C. Justice, Xinyu Zhang, Zuoheng Wang, Dana B. Hancock, Eric O. Johnson, Ke Xu

## Abstract

**Background:** Cocaine use accelerates human immunodeficiency virus (HIV) progression and worsens HIV outcomes. We assessed whether DNA methylation in blood mediates the association between cocaine use and HIV severity in a veteran population.

**Methods:** We analyzed 1,435 HIV-positive participants from the Veterans Aging Cohort Study Biomarker Cohort (VACS-BC). HIV severity was measured by the Veteran Aging Cohort Study (VACS) index. We assessed the effect of cocaine use on VACS index and mortality among the HIV-positive participants. We selected candidate mediators that were associated with both persistent cocaine use and VACS index by epigenome-wide association (EWA) scans at a liberal p-value cutoff of 0.001. Mediation analysis of the candidate CpG sites between cocaine’s effect and the VACS index was conducted, and the joint mediation effect of multiple CpGs was estimated. A two-step epigenetic Mendelian randomization (MR) analysis was conducted as validation.

**Results:** More frequent cocaine use was significantly associated with a higher VACS index (β=1.00, p=2.7E-04), and cocaine use increased the risk of 10-year mortality (hazard ratio=1.10, p=0.011) with adjustment for confounding factors. Fifteen candidate mediator CpGs were selected from the EWA scan. Twelve of these CpGs showed significant mediation effects, with each explaining 11.3%-29.5% of the variation. The mediation effects for 3 of the 12 CpGs were validated by the two-step epigenetic MR analysis. The joint mediation effect of the 12 CpGs accounted for 47.2% of cocaine’s effect on HIV severity. Genes harboring these 12 CpGs are involved in the antiviral response *(IFIT3, IFITM1, NLRC5, PLSCR1, PARP9)* and HIV progression *(CX3CR1, MX1)*.

**Conclusions:** We identified 12 DNA methylation CpG sites that appear to play a mediation role in the association between cocaine use and HIV severity.

## Introduction

Cocaine use is common among persons with chronic human immunodeficiency virus (HIV) infection, with prevalence estimates for current or recent use ranging from 5% to 30% [1-6], compared with 2% in the US general population [7]. Previous studies have shown that cocaine use accelerated HIV progression [8-11]. However, the biological mechanism of cocaine’s effect on HIV outcomes remains largely unknown. Some studies have suggested that cocaine use may worsen HIV outcomes due to poor adherence to antiretroviral therapy (ART) among HIV-positive participants [2, 12]. Other studies have demonstrated that cocaine’s adverse effect on HIV outcomes is independent of ART [10, 11, 13-15], supporting the hypothesis that cocaine exposure may lead to long-lasting pathophysiological changes in the immune system that worsen HIV outcomes.

DNA methylation (DNAm) is an important mechanism associated with many environmental exposures such as smoking, alcohol, and drug misuse [16-26] and diseases such as cancer, diabetes and cardiovascular diseases [27-32]. Our previous study showed that two DNAm CpG sites in *NLRC5* were differentially methylated between HIV-positive and HIV-negative participants in peripheral blood [33]. DNAm may play an important mediation role linking environmental exposure and disease outcomes [34-40]. Environmental exposure such as substance use or toxicants can directly or indirectly affect DNA methylatransferases, causing global or site-specific DNAm changes that may lead to disease [41]. A recent study reported that DNAm sites in *PIM3* (energy metabolism) and *ABCG1* (lipid metabolism) mediated the association between prenatal famine exposure and long-term metabolic outcomes [38]. Another study reported the mediation effect of cg05575921 *(AHRR)* between smoking and the risk of bladder cancer among postmenopausal women [42].

Previous studies have shown that the use of cocaine enhances HIV-1 replication and undermines immune function by dysregulating gene expression on HIV-1 entry co-receptors, enhancing HIV-1 cellular toxicity and dysregulating interleukins (IL) in the host [43, 44]. Cocaine use increases the release of cytokines in immune cells and alters cytokine profile in HIV-infected individuals [45, 46]. Specifically, cocaine use was positively associated with IL-4 and IL-10 [47], which likely worsens HIV severity and disease progression. Epigenetic mechanisms may play a role in cocaine’s effect on the HIV severity because cocaine exposure has been showed to increase the expression of Methyl CpG binding protein 2 (MeCP2) expression [48] as well as *DNMT3A* and *DNMT3B* expressions [49] in the animal brains. In a well-matched, case-control human pilot study, cocaine use alters DNA methylation profile in blood [50]. It is plausible that cocaine use may lead to DNAm changes in immune response genes and gene expression changes in cytokine gene family, which further affects HIV progression. Thus, we hypothesized that DNAm may mediate the effect of cocaine exposure on HIV severity.

In this study, we first validated previous findings by examining cocaine’s adverse effect on HIV severity and mortality. We further conducted mediation analyses to assess the mediation role of DNAm sites (or CpGs) on cocaine’s effect on HIV severity using the Veteran Aging Cohort Study Biomarker Cohort (VACS-BC, n=1,435). To assess how sensitive our results are to the violation of model assumptions and validate our findings using a different approach, we performed the sensitivity analysis [51] and the two-step epigenetic Mendelian randomization (MR) analysis that used genetic variants as instrumental variables to assess the mediation role of DNAm between cocaine use and HIV severity [52]. Our results provide new insights for the role of DNAm on how cocaine affects HIV severity.

## Methods

### Study samples

VACS is a prospective cohort study of veterans designed to study substance use and HIV-related outcomes with patient surveys, electronic medical records and biospecimen data [53]. A baseline survey was conducted at enrollment [53]. The follow-up survey of 5 visits occurred at approximately one-year intervals [53]. Blood samples were collected in the middle of follow-up for a subset of participants in the cohort (VACS-BC) [54]. A total of 1,435 HIV-positive participants from the VACS-BC were used to examine cocaine’s effect on mortality and HIV severity, and a subset of participants (n=875) with DNAm data available were used for mediation analyses (**Figure 1**). Demographic and clinical information of baseline samples and a subset of the samples at the time of blood collection are summarized in **Table 1**.

**Figure 1.**
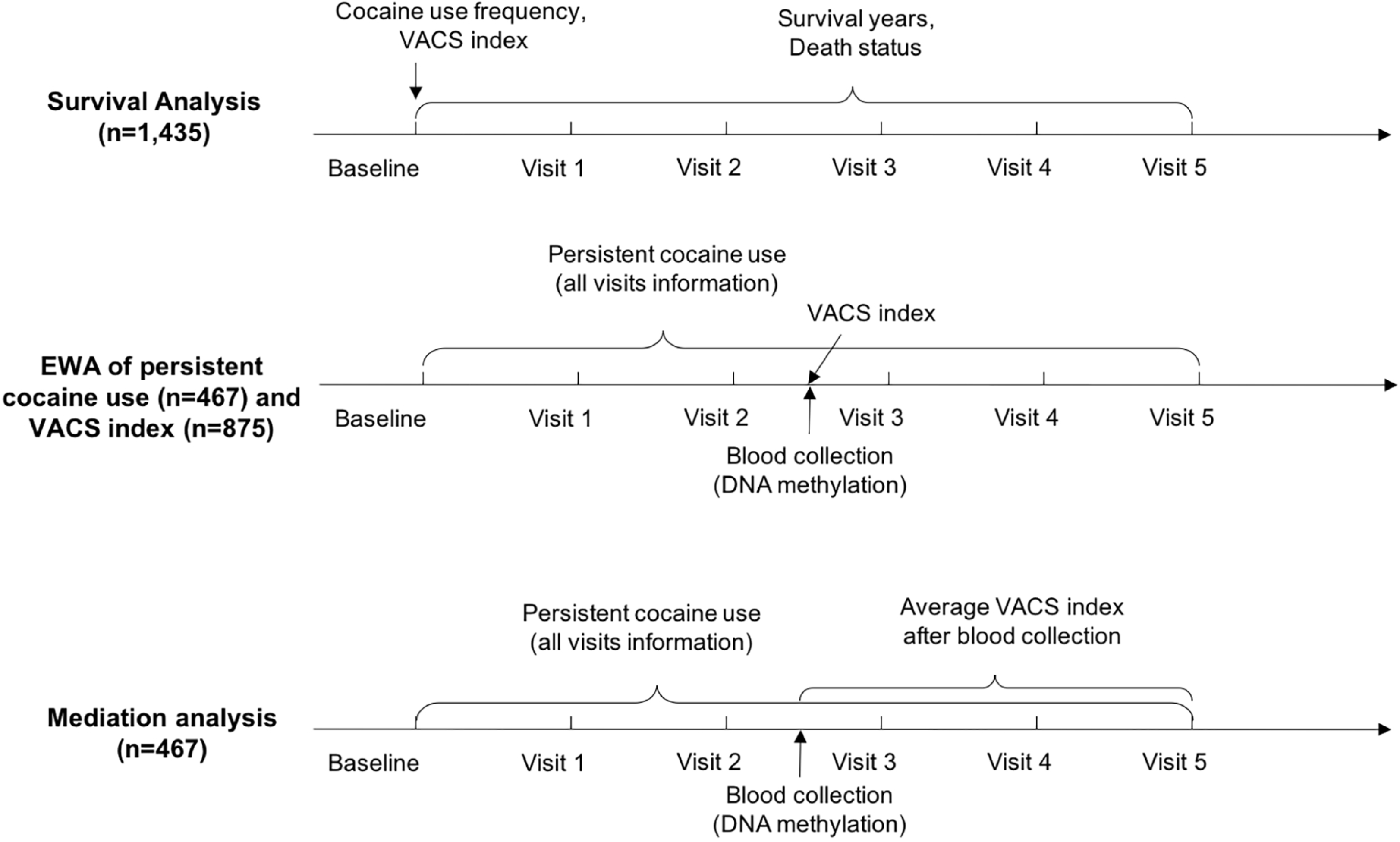
Timeline of data and blood sample collection for each analysis

**Table 1:**
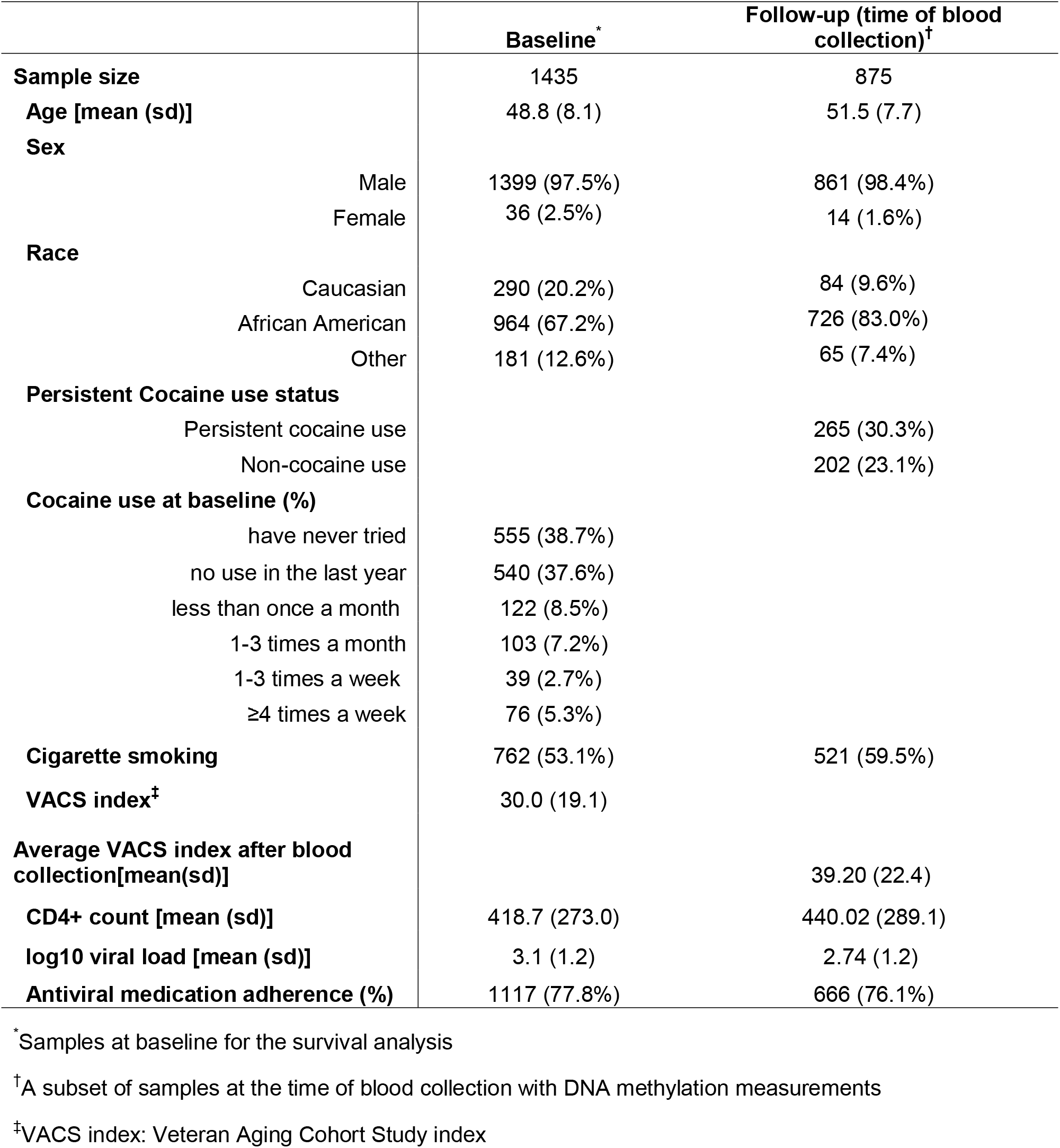
Sample characteristics in HIV-positive participants.

|  | Baseline<sup>*</sup> | Follow-up (time of blood collection)<sup>†</sup> |
| --- | --- | --- |
| **Sample size** | 1435 | 875 |
| **Age [mean (sd)]** | 48.8 (8.1) | 51.5 (7.7) |
| **Sex** |  |  |
| Male | 1399 (97.5%) | 861 (98.4%) |
| Female | 36 (2.5%) | 14 (1.6%) |
| **Race** |  |  |
| Caucasian | 290 (20.2%) | 84 (9.6%) |
| African American | 964 (67.2%) | 726 (83.0%) |
| Other | 181 (12.6%) | 65 (7.4%) |
| **Persistent Cocaine use status** |  |  |
| Persistent cocaine use |  | 265 (30.3%) |
| Non-cocaine use |  | 202 (23.1%) |
| **Cocaine use at baseline (%)** |  |  |
| have never tried | 555 (38.7%) |  |
| no use in the last year | 540 (37.6%) |  |
| less than once a month | 122 (8.5%) |  |
| 1-3 times a month | 103 (7.2%) |  |
| 1-3 times a week | 39 (2.7%) |  |
| ≥4 times a week | 76 (5.3%) |  |
| **Cigarette smoking** | 762 (53.1%) | 521 (59.5%) |
| **VACS index<sup>‡</sup>** | 30.0 (19.1) |  |
| **Average VACS index after blood collection[mean(sd)]** |  | 39.20 (22.4) |
| **CD4+ count [mean (sd)]** | 418.7 (273.0) | 440.02 (289.1) |
| **log10 viral load [mean (sd)]** | 3.1 (1.2) | 2.74 (1.2) |
| **Antiviral medication adherence (%)** | 1117 (77.8%) | 666 (76.1%) |

<sup>\*</sup>Samples at baseline for the survival analysis
<sup>†</sup>A subset of samples at the time of blood collection with DNA methylation measurements
<sup>‡</sup>VACS index: Veteran Aging Cohort Study index

### Assessment of cocaine use

The timeline of cocaine use assessment for each analysis is illustrated in **Figure 1**. Information on cocaine use status was self-reported through telephone interviews for a total of 5 visits. We defined the “persistent cocaine use” group as self-reported cocaine use across all 5 visits and the “no cocaine use” group as self-reported no cocaine use across all 5 visits. This definition led to a subset of samples with 265 persistent cocaine users and 202 nonusers for the mediation analyses to eliminate the inconsistent response across 5 visits and examine the effect of long-term cocaine exposure on DNAm and HIV severity.

The frequency of cocaine use was also assessed at baseline (**Figure 1**). Each participant was asked “how often in the past year have you used cocaine or crack?”, from which cocaine frequency of use was coded as an ordinal variable as follows: 0=never tried; 1= no use in the last year; 2=less than once a month; 3=1-3 times a month; 4=1-3 times a week; 5=4 or more times a week.

### Assessment of mortality and HIV severity

The timeline of HIV severity measurement and survival information for each analysis is shown in **Figure 1**. Mortality and survival year information were based on medical records. The VACS index was used as a measure of HIV severity [55-58] and was obtained at each visit and at the time of blood collection (**Figure 1**). The VACS index was calculated by summing preassigned points for age, routinely monitored indicators of HIV disease (CD4 count and HIV-1 RNA) and other general indicators of organ system injury [55]. A high VACS index corresponds to worsened HIV outcomes, and the VACS index is positively associated with increased mortality [59]. The VACS index and DNAm profiling were measured at the same time for the selection of candidate mediator CpGs, and the average VACS index after blood collection was used for mediation analyses (**Figure 1**).

### DNA methylation profiling and quality control

DNA samples were extracted from blood for a subset of 875 HIV-positive participants (**Figure 1**). DNAm was profiled using two different methylation arrays, with 475 samples profiled by the Infinium Human Methylation 450K BeadChip (HM450K, Illumina Inc., CA, USA) and 400 samples later profiled by the Infinium Human Methylation EPIC BeadChip (EPIC, Illumina Inc., CA, USA) [54]. DNA samples were randomly selected for each methylation array regardless of cocaine use status or other clinical demographic variables.

The quality control (QC) for samples measured by each array was conducted separately using the same pipeline as previously described [60] by the R package *minfi* [61]. After QC, a total of 408,583 CpGs measured by both the HM450k and EPIC array remained for analysis. Six cell type proportions (CD4+ T cells, CD8+ T cells, NK T cells, B cells, monocytes and granulocytes) were estimated for each participant using the established method [62]. Negative control probes were designed to capture background signals in Illumina arrays, and negative control principal components (PCs) were extracted by *minfi* to control for background noise [61]. Batch effect removal was conducted by *combat* after QC [63].

### Genotyping and quality control

The 1,177 samples were genotyped using the Illumina HumanOmniExpress Beadchip and imputed for 18,960,156 single nucleotide polymorphisms (SNPs). *IMUPTE2* (ver 2.3.2) was used for imputation with the reference of 1000 genome phase 3 [64]. QC was conducted using *plink* (ver 1.90b21) [65]. SNPs and samples with low call rate less than 0.05 were removed. The Hardy-Weinberg equilibrium test cutoff was set to 1E-06. SNPs with minor allele frequency less than 0.01 were filtered.

## Statistical analysis

### Cocaine survival analysis among HIV-positive participants at baseline

Survival analysis was conducted using baseline information among 1,435 HIV-positive participants with cocaine use frequency (0-5) and other covariates (**Figure 1**). Kaplan-Meier analyses on 10-year follow-up among HIV-positive and HIV-negative participants by cocaine use frequency (0-5) at baseline were conducted, and the Kaplan-Meier curves were plotted by using the R package *survminer* [66]. A test on ordered differences of Kaplan-Meier curves by cocaine use frequency was conducted by *survminer* [66].

To adjust for confounding factors, a Cox proportional-hazards model was used to assess the hazard ratio of baseline cocaine use frequency (0-5) on mortality during the follow-up using the R package *survival* [67]. The following model was used to calculate the adjusted hazard ratio among HIV-positive participants:

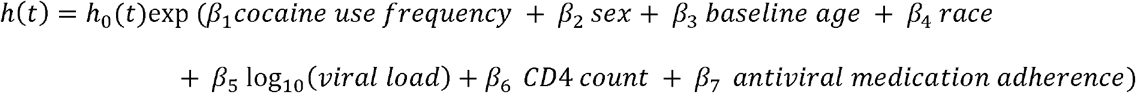

### Association between cocaine use frequency and HIV severity among HIV-positive participants at baseline

This analysis was conducted using baseline information on cocaine use frequency (0**-**5), the VACS index and other covariates (**Figure 1**). The following linear regression model was performed to test the association of cocaine use frequency and HIV severity, adjusting for confounders as shown in the following model:

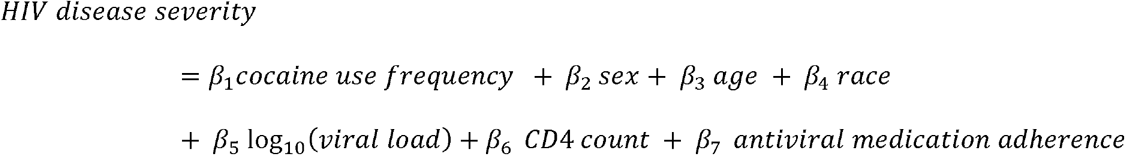

### Selection of candidate CpGs by epigenome-wide association (EWA) of persistent cocaine use and HIV severity

To select candidate CpGs for mediation analysis, we conducted two separate EWAs, one for persistent cocaine use and the other for HIV severity (**Figure 1**). Each EWA model adjusted for sex, baseline age, race, smoking, self-reported antiviral medication adherence, white blood cell count, estimated cell type proportions and negative control PCs. We used the linear regression model with methylation as dependent variable for EWA as described previously [33, 60, 68]. Since CD4+ T cell count is one component of the VACS index, to avoid overrepresented CpGs associated with CD4+ T cells in the EWA results, we extracted the top 1,000 CD4+ T cell type relevant CpGs based on data from *FlowSorted.Blood.450k* [69]. The top 2 PCs that in total account for > 80% variation of the 1,000 CD4+ T cell CpGs were used as covariates in the VACS index EWA model. CpGs with p<0.001 in both EWAs for persistent cocaine use and HIV severity were selected as candidate CpGs for mediation analyses. A liberal selection threshold was arbitrarily set to make sure there would be a sufficient number of candidate CpGs for the mediation analysis. To limit confounding by use of other substances, we tested the association of each candidate CpG site with alcohol use, cannabis use, and opioid use based on self-reported data. Alcohol use was assessed by using 3 items of Alcohol Use Diagnosis Identification Test-consumption (AUDIT-C). Cannabis and opioid uses were assessed by asking the same questions as for cocaine use, described earlier.

### Single-site mediation analysis and joint mediation analysis

The selected candidate CpGs were assessed as potential mediators of the association between persistent cocaine use and HIV severity among HIV-positive participants (n=467). We performed single-site mediation analysis using the mediation method as previously described [51] and the R package *mediation* [70]. Here, we used the average VACS index after DNAm profiling to ensure the temporality of our mediation hypothesis that DNAm measurement preceded the HIV severity measurement. In our mediation model, we adjusted for sex, age, race, smoking, self-reported antiviral medication adherence, white blood cell count, and estimated cell type proportions as confounding factors.

We used *M* to represent the candidate CpGs (mediator), *X* to represent persistent cocaine use status (exposure), *Y* to represent the average VACS index after blood collection (outcome), and *C_t_* to represent *k* confounding variables (sex, age, race, smoking, self-report antiviral medication adherence, white blood cell count, estimated CD8 T cells, granulocytes, NK cells, B cells, and monocytes). The mediator mode *f*(*M*|*X*, *C*) examined the association between persistent cocaine use and CpGs:

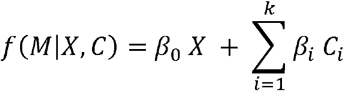

The outcome model *f(Y|X, M, C)* examined both the direct effect of persistent cocaine use on VACS index and the mediation effect by CpG:

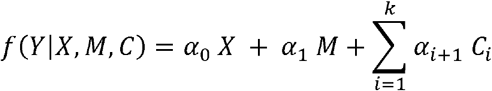

Thus, the mediation effect, or the average causal mediation effect (ACME) of CpG *M* is *α*_1_*β*_0_, the total effect is *α*_0_ *+ α*_1_*β*_0_, and the proportion mediated is *α*_1_*β*_0_/(*α*_0_ *+ α*_1_*β*_0_) The confidence interval and p-value were estimated by bootstrapping 1,000,000 iterations.

To assess the robustness of the results if the sequential ignorability assumption was violated, we conducted a sensitivity analysis developed by Imai et al. [51] using the R package *mediation* [70]. Sequential ignorability consists of two assumptions: (a) conditional on the covariates *C_i_*, the exposure *X* is independent of all potential values of the outcome *Y* and mediator *M;* and (b) the observed mediator *M* is independent of all potential outcomes *Y* given the observed exposure *X* and covariates *C_i_*. The sensitivity parameter ρ was calculated on a grid of 0.05 and he ρ at which ACME = 0 was calculated. For each mediator, sensitivity plots were illustrated to show the estimated ACME and their 95% confidence interval as a function of ρ (**Figure S2**). If the ρ at which ACME = 0 is close to 0, it indicates that the mediation analysis was sensitive to violation of the sequential ignorability assumption.

The joint mediation analysis of all significant mediator CpGs were conducted as previously described [71]. The mediator model *f(M_j_|X, C)* for multiple mediators *M_j_* (*M*_1_ *M*_2_,…, *M_n_*) is:

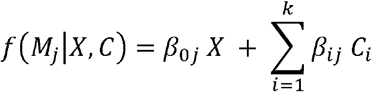

The outcome model *f*(*Y*|*X, M*_1_,…, *M_n_*, *C*) is:

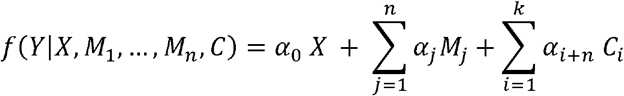

The joint mediation effect of CpGs *M*_1_, *…, M_j_* is 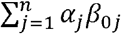, the total effect is 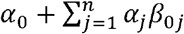, and the proportion mediated is 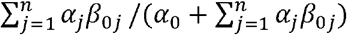. The confidence interval and p-value were estimated by bootstrapping 1,000,000 iterations.

### Two-step epigenetic Mendelian randomization of cocaine and HIV severity

To evaluate whether the results from the mediation analysis were influenced by reverse causation or unmeasured confounding, we conducted a two-step epigenetic MR analysis [52] (n=1,177) on cocaine use, candidate mediator CpGs, and HIV severity using the inverse-variance weighted (IVW) method by R package *MendelianRandomization* [72].

In step 1, we conducted a two-sample MR on the effect of cocaine use on candidate CpGs (n=1,177). Based on a recent meta-analysis of a cocaine dependence genome-wide association study (GWAS) [73], 8 SNPs genotyped in our samples pruned at linkage disequilibrium (LD) r^2^<0.1 by the R package *LDlinkR* [74] were used as instrumental variables (p<1E-05) (**Table S4**). We tested the associations between the 8 SNPs and the candidate CpGs, adjusting for age, sex, race and 5 ancestry PCs using linear regression model in our sample (n=1,177). Based on these summary statistics, we conducted MR using the IVW method to evaluate the effect of cocaine use on candidate CpGs.

In step 2, we conducted a one-sample MR on the effect of candidate CpGs on HIV severity (n=1,177). Here, *cis-* methylation quantitative trait loci (meQTLs) were used as instrumental variables. cis-meQTLs were defined by the distance between a candidate CpG and a SNP within 1 Mb. A linear regression analysis was performed to identify cis-meQTLs, adjusted for age, sex, race and 5 ancestry PCs. For each candidate CpG, cis-meQTLs with p<0.01 after pruning (LD r^2^<0.1 using 1000 genome African ancestry samples as references [75]) were used as instrumental variables in the MR analysis (**Table S4**) by the R package *LDlinkR* [74]. Association between each cis-meQTL and HIV severity was assessed by linear regression, adjusting for age, sex and 5 ancestry PCs. Similar to the first step, we conducted an MR using the IVW method to evaluate the effect of candidate CpGs on HIV severity.

## Results

### Cocaine use affects HIV severity and mortality among HIV-positive participants

We found that among HIV-positive participants, higher cocaine use frequency was associated with increased mortality (p=0.008, **Figure 2a**). This difference was not found among HIV-negative participants (p= 0.180, **Figure 2b**). Using Cox proportional-hazards model, this trend remained significant with a hazard ratio (HR) of 1.10 (95% CI: 1.02-1.19, p=0.011), controlling for sex, baseline age, race, viral load, CD4 count and antiviral medication adherence (**Table 2**). A higher frequency of cocaine use at baseline was also significantly associated with a higher VACS index (i.e., higher HIV severity, (β=1.00, p= 0.00027) after adjusting for sex, age, race, viral load, CD4 count and antiviral medication adherence (**Table 2**). To account for other drug use, we further adjusted for baseline use of alcohol, cigarette smoking, cannabis, and opioids in the model. Cocaine use frequency remains significantly associated with HIV severity after adjusting for use of other substances (p=0.049). Our results suggest that cocaine use accelerated HIV progression and increased mortality independent of antiviral medication adherence, which is consistent with previous reports [10, 11, 13-15].

**Figure 2.**
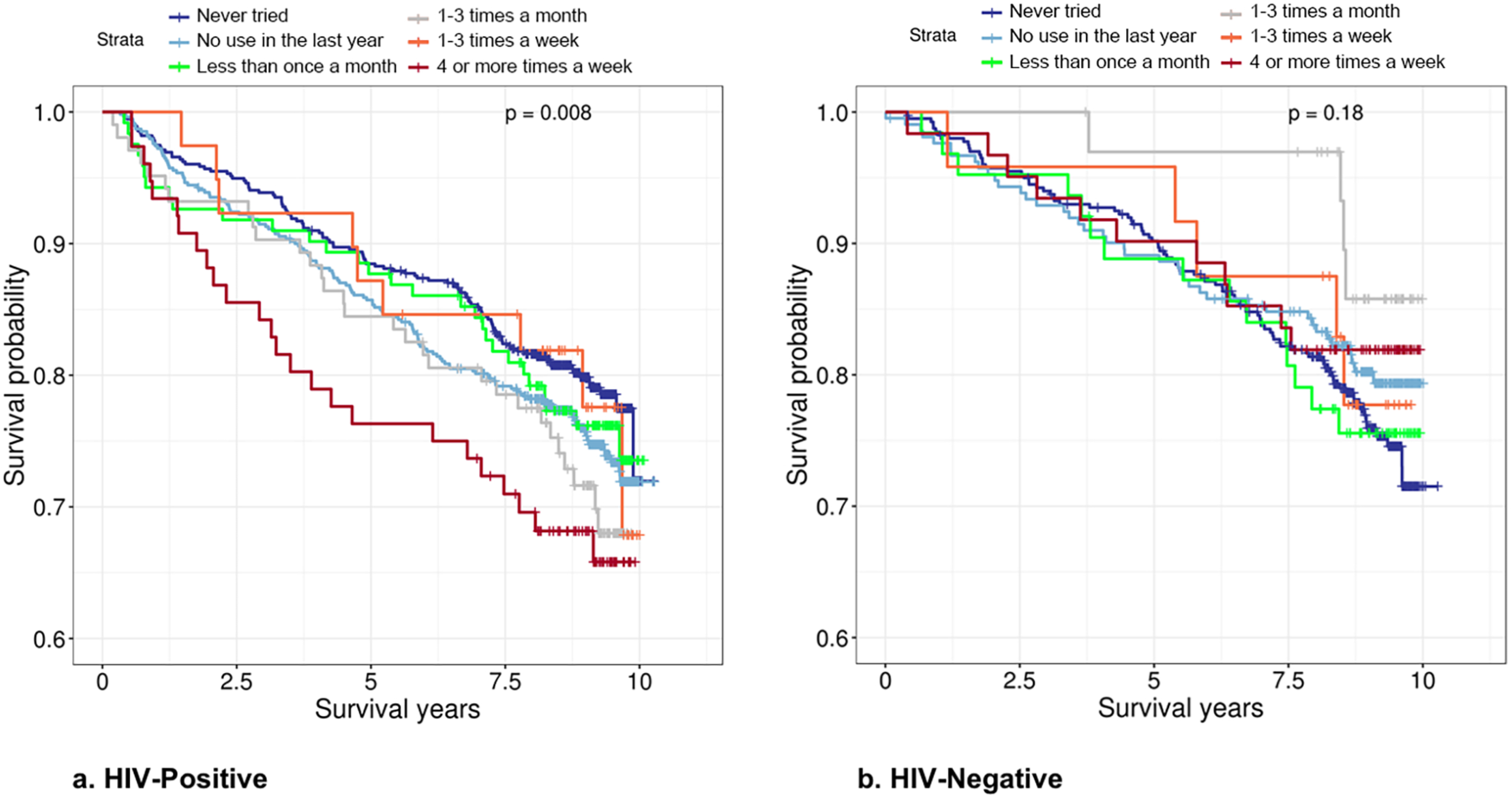
Kaplan-Meier curves of cocaine use frequency at baseline among HIV-positive (n=1,435, **2a**) and HIV-negative (n=795, **2b**) participants. The higher frequency of cocaine use is associated with lower survival probability among HIV-positive participants but not among HIV-negative participants.

**Table 2:** Association between cocaine use frequency and HIV severity and survival analysis among HIV-positive participants (n=1,435)

|  | HIV severity<sup>*</sup> |  |  |  | Mortality<sup>†</sup> |  |  |  |
| --- | --- | --- | --- | --- | --- | --- | --- | --- |
|  | Estimate | SE | T | P value | Hazard ratio | 95% CI | Z | P value |
| Cocaine use frequency<sup>‡</sup> | 1.00 | 0.28 | 3.65 | 2.70E-04 | 1.10 | (1.02,1.19) | 2.54 | 1.10E-02 |
| Sex (reference: male) | 5.78 | 2.33 | 2.48 | 1.34E-02 | 0.27 | (0.07,1.08) | -1.86 | 6.33E-02 |
| Age<sup>‡</sup> | 1.09 | 0.05 | 23.58 | < 2E-16 | 1.05 | (1.04,1.07) | 7.35 | 1.90E-13 |
| Race (reference: Caucasian) |  |  |  |  |  |  |  |  |
| African American | 7.30 | 0.94 | 7.73 | 2.00E-14 | 1.05 | (0.80,1.39) | 0.37 | 7.15E-01 |
| Other | 5.33 | 1.31 | 4.08 | 4.80E-05 | 0.71 | (0.47,1.09) | -1.57 | 1.17E-01 |
| log 10 viral load<sup>‡</sup> | 4.72 | 0.36 | 13.11 | < 2eE-16 | 1.07 | 2 | 1.36 | 1.73E-01 |
| CD4 count<sup>‡</sup> | -0.03 | 0.00 | -19.05 | < 2E-16 | 1.00 | (1.00,1.00) | -3.74 | 1.80E-04 |
| Antiviral medication adherence<sup>‡</sup> | 2.59 | 0.99 | 2.62 | 8.87E-03 | 1.02 | (0.76,1.37) | 0.13 | 8.95E-01 |

<sup>\*</sup>Summary statistics from linear regression model
<sup>†</sup>Summary statistics from Cox proportional hazards model
<sup>‡</sup>Measured at baseline

### Selection of candidate DNAm sites for mediation analysis by EWA scan

The EWA scan of persistent cocaine use showed good control of inflation (λ=1.034, **Figure S1**). A total of 497 CpGs met our candidate selection threshold (p<0.001). The top ranked CpG site, cg22917487, was close to the epigenome-wide significance threshold with p-value of 1.69E-07. This CpG site is located in *CX3CR1*, a gene that encodes a coreceptor for HIV-1 and leads to rapid HIV progression (**Table S1**).

The EWA scan of the VACS index also showed good control of inflation (λ=1.116, **Figure S1**). There were 876 CpGs that reached the candidate selection threshold (p<0.001) (**Table S2**). Of note, 6 CpGs reached the epigenome-wide significance threshold (p<1.2E-07). These CpGs were located near the genes involved in the viral and immune response *(PARP9, IFITM1, CD247, IFIT3, VASN*, and *RUNX1)*.

We selected candidate CpGs that were both associated with cocaine use and HIV severity (p<0.001) by two separate EWA scans of 408,583 CpGs for mediation analysis. Fourteen CpGs met both candidate selection thresholds. Additionally, cg22917487 in *CX3CR1* showed a strong association with cocaine (p=1.69E-07) and a marginal association with the VACS index (p=1.73E-03). Given its biological plausibility, this CpG was also included as a candidate mediator for mediation analysis. Five of the top 10 VACS index EWA CpGs were selected as candidate mediator CpGs (cg08122652, *PARP9*, p=2.30E-10; cg03038262, *IFITM1*, p=7.65E-09; cg06188083, *IFIT3*, p=4.76E-08; cg08818207, *TAP1*, p=2.11E-07; cg26312951, *MX1*, p=2.50E-07). Overall, a total of 15 CpGs were selected as candidates to assess their potential mediation roles on the association between persistent cocaine use and HIV severity. Notably, the DNAm from each of the 15 CpGs was not associated with cannabis, opioid or alcohol use (p>0.05, **Table S3**).

### Mediation analysis of candidate CpGs between persistent cocaine use and HIV severity

We examined the mediation role of DNAm between persistent cocaine use and HIV severity. Twelve out of the 15 candidate CpGs showed significant mediation effects on the association between persistent cocaine use and the VACS index, with p-values ranging from 1.00E-06 to 0.00307 (**Table 4**). These results remained significant after Bonferroni correction (p<0.0033). Each CpG mediator explained between 11.3% to 29.5% of persistent cocaine use affecting HIV severity. Notably, the direction of mediation effects among these 12 mediator CpGs were the same. The average direct effects of cocaine on HIV severity were attenuated from 0.329 to 0.231-0.291 after adjusting for each mediator CpG. These 12 CpGs collectively mediated 47.2% of the cocaine’s effects on HIV severity by joint mediation analysis.

We also conducted a sensitivity analysis on these 15 candidate CpGs to assess the robustness of our mediation analysis when the sequential ignorability assumption was violated [51]. The absolute sensitivity parameters at which ACME=0 of the 12 significant mediator CpGs were relatively higher (|ρ|≥0.15) than 3 nonsignificant CpGs (|ρ|≤0.10) (**Table 4**, **Figure S2**). Notably, 6 significant mediator CpGs had |ρ| of 0.30, indicating that these mediation effects were robust even when the assumptions are slightly violated. The sensitivity analysis showed that our mediation results were relatively stable.

Significant mediator CpGs are located near 11 viral and immune response genes: *MX1, PARP9, IFIT3, IFITM1, NLRC5, EPSTI1, PLSCR1, TAP2, TAP1, CX3CR1* and *RIN2*. Five CpGs are located on 5’ gene regulatory regions, 4 CpGs on gene bodies, 2 CpGs on transcription start sites and 1 CpG on 3’ gene regulatory region. Notably, these 12 CpGs were mostly less methylated in the persistent cocaine use group than in the no cocaine use group (**Table 3**, **Figure 3**). **Figure 4** illustrates the mediation effect of cg26312951 (MX1), cg08122652 *(PARP9)*, cg07839457 *(NLRC5)* and cg22917487 *(CX3CR1)* on persistent cocaine use affecting HIV severity.

**Table 3:** The selected candidate CpG sites by epigenome-wide association (EWA) scan on persistent cocaine use (n=467) and HIV severity (n=875)

| CpG | Chr | Position | Nearest gene | Cocaine EWA effect size | Cocaine EWA p-value | VACS index* EWA effect size | VACS index* EWA p-value | Reference Gene Group | Relations to CpG Islands |
| --- | --- | --- | --- | --- | --- | --- | --- | --- | --- |
| cg22917487 | 3 | 39322103 | *CX3CR1* | 1.71E-02 | 1.69E-07 | 1.96E-04 | 1.73E-03 | Body;TSS200;5UTR;TSS1500 |  |
| cg07839457 | 16 | 57023022 | *NLRC5* | -5.18E-02 | 3.16E-05 | -7.08E-04 | 4.02E-04 | TSS1500 | N_Shore |
| cg26312951 | 21 | 42797847 | *MX1* | -3.93E-02 | 3.24E-05 | -7.80E-04 | 2.50E-07 | TSS200;5UTR | N_Shore |
| cg06188083 | 10 | 91093005 | *IFIT3* | -5.06E-02 | 3.34E-05 | -9.96E-04 | 4.76E-08 | Body |  |
| cg08122652 | 3 | 1.22E+08 | *PARP9* | -6.13E-02 | 1.10E-04 | -1.26E-03 | 2.30E-10 | 5UTR;TSS1500 | N_Shore |
| cg22385827 | 2 | 2.11E+08 | *C2orf67* | 8.10E-03 | 2.01E-04 | 1.20E-04 | 7.52E-04 | TSS1500 | Island |
| cg22930808 | 3 | 1.22E+08 | *PARP9* | -6.52E-02 | 2.44E-04 | -1.17E-03 | 1.52E-06 | 5UTR;TSS1500 | N_Shore |
| cg06981309 | 3 | 1.46E+08 | *PLSCR1* | -4.29E-02 | 2.63E-04 | -5.72E-04 | 4.21E-04 | 5UTR | N_Shore |
| cg03753191 | 13 | 43566902 | *EPSTI1* | -1.36E-02 | 2.81E-04 | -2.44E-04 | 5.50E-05 | TSS1500 | S_Shore |
| cg00096307 | 2 | 1.07E+08 | *UXS1* | -1.12E-02 | 3.59E-04 | 1.74E-04 | 5.73E-04 | Body |  |
| cg26396492 | 20 | 19915762 | *RIN2* | -1.26E-02 | 4.76E-04 | -2.00E-04 | 9.15E-04 | Body |  |
| cg22940798 | 6 | 32805554 | *TAP2* | -1.93E-02 | 4.82E-04 | -2.89E-04 | 4.72E-04 | Body | N_Shore |
| cg08623256 | 5 | 3858275 |  | -1.73E-02 | 6.96E-04 | 3.17E-04 | 7.18E-04 |  | S_Shelf |
| cg08818207 | 6 | 32820355 | *TAP1* | -2.67E-02 | 8.18E-04 | -6.18E-04 | 2.11E-07 | Body | N_Shore |
| cg03038262 | 11 | 315262 | *IFITM1* | -2.98E-02 | 9.59E-04 | -6.46E-04 | 7.65E-09 | 3UTR | N_Shore |

\*VACS index: Veteran Aging Cohort Study index

**Table 4:**
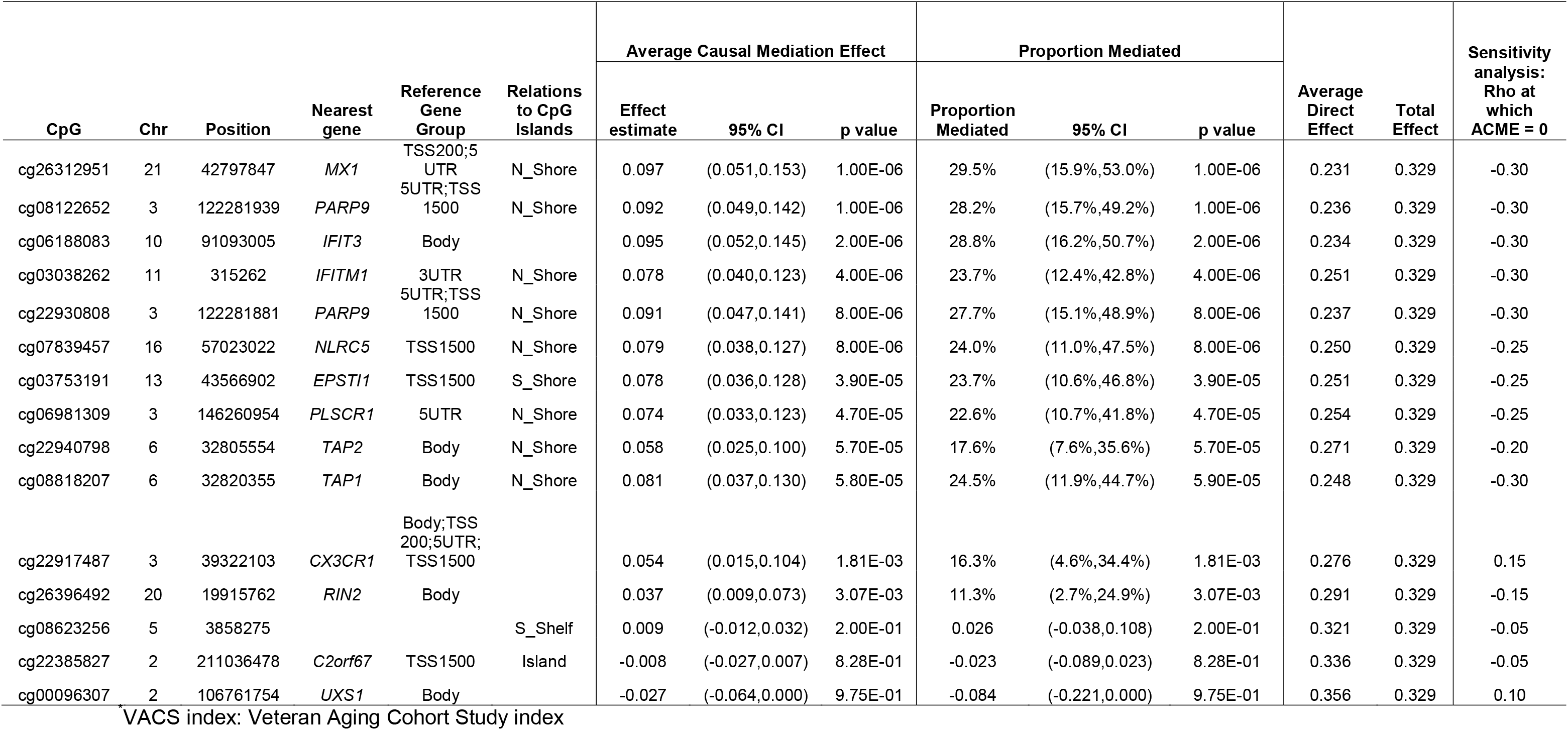
Mediation analyses on candidate CpGs between cocaine use and VACS index^*^ (n=467)

**Figure 3.**
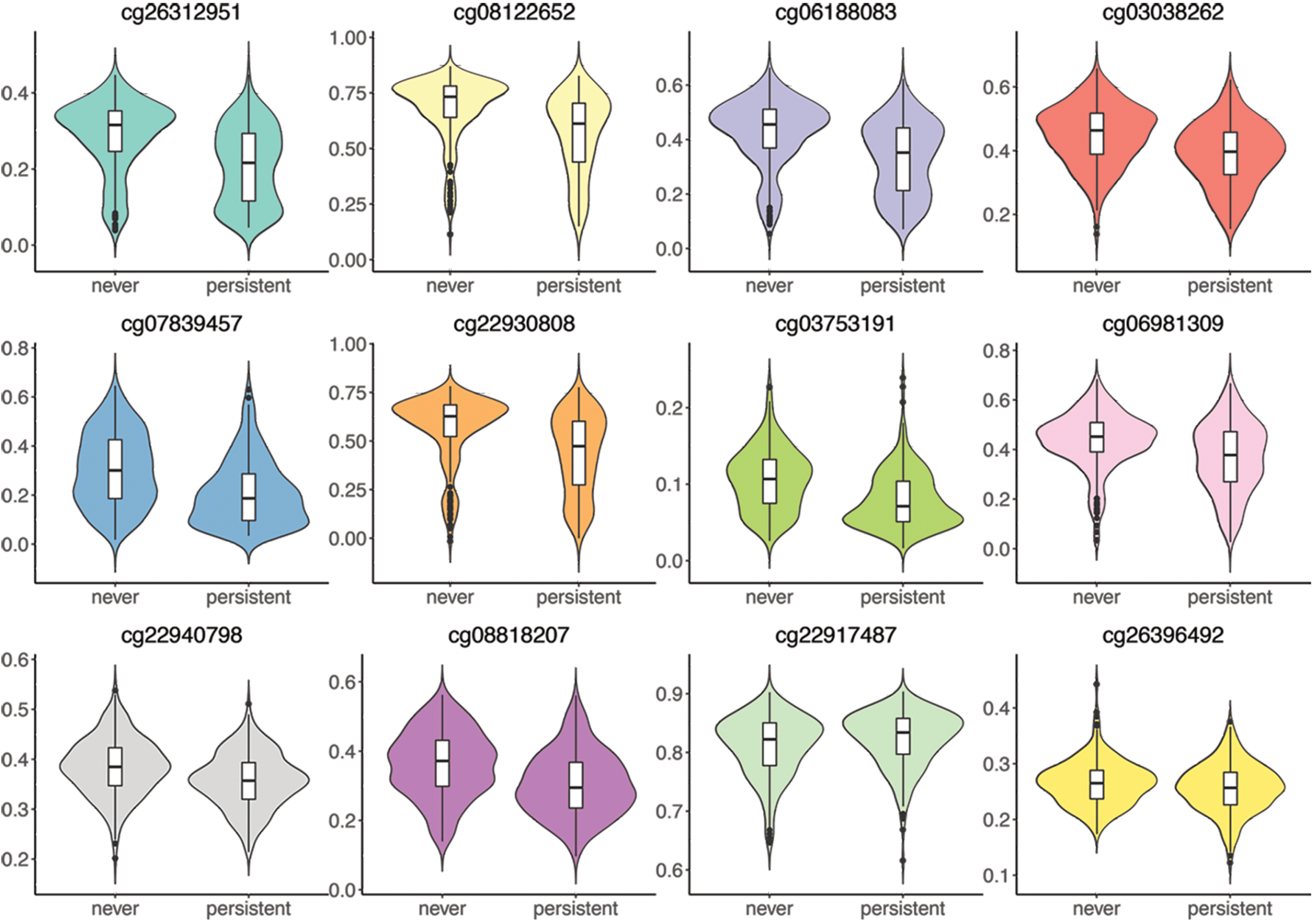
DNA methylation level of the significant CpG mediators by persistent cocaine use status

**Figure 4.**
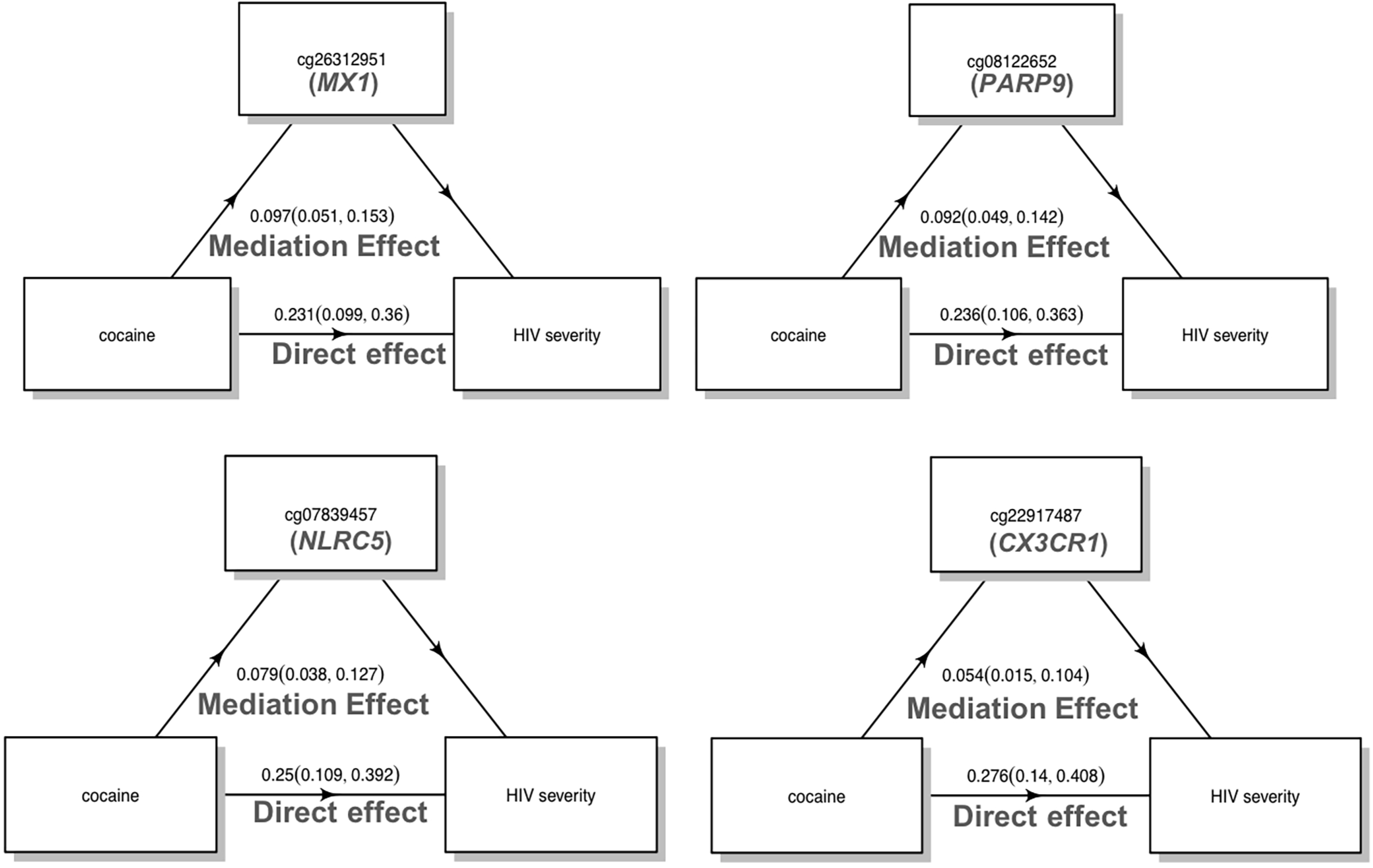
Significant mediation effect of cg26312951 *(MX1)*, cg08122652(PARP9), cg07839457 *(NLRC5)*, and cg22917487 *(CX3CR1)* between persistent cocaine use and HIV severity (p<0.0033)

### Two-step epigenetic Mendelian randomization of cocaine and HIV severity

To validate our mediation results while eliminating unmeasured confounding and reverse causation, we used the two-step epigenetic MR method [52] to test our mediation hypotheses (n=1,177): whether cocaine use has a causal effect on candidate CpGs (step 1) and whether candidate CpGs have causal effects on HIV severity (step 2).

In step 1, we conducted the MR analysis based on summary statistics of a meta-analysis of GWAS on cocaine dependence [73]. The effect estimates of the association between 8 SNP instrumental variables and 15 candidate CpG sites were obtained in our sample. Our MR analysis showed that cocaine had significant MR estimates (p<0.05) on 4 CpGs (cg03753191, *EPSTI1;* cg06188083, *IFIT3;* cg26312951,*MX1*; cg22917487,*CX3CR1)*, as shown in **Table 5**. Three of these CpGs were also among the top significant mediators in our previous mediation analysis (**Table 4**).

**Table 5:** Two-step epigenetic mendelian randomization on cocaine and HIV severity (n=1,177)

| CpG | Chr | Position | Nearest gene | Step 1: cocaine affecting Methylation |  |  |  |  | Step 2: methylation affecting HIV severity |  |  |  |  |
| --- | --- | --- | --- | --- | --- | --- | --- | --- | --- | --- | --- | --- | --- |
|  |  |  |  | Numb<br>er of<br>IV* | Estimate | Standard<br>Error | 95% CI | p value<sup>†</sup> | Numb<br>er of<br>IV* | Estimate | Standard<br>Error | 95% CI | p value<sup>†</sup> |
| cg03753191 | 13 | 43566902 | *EPSTI1* | 8 | 0.206 | 0.070 | (0.069,0.343) | **3.27E-03** | 9 | -10.688 | 2.300 | (-15.197,-6.180) | **3.38E-06** |
| cg06188083 | 10 | 91093005 | *IFIT3* | 8 | 0.269 | 0.109 | (0.056,0.482) | **1.33E-02** | 4 | -6.756 | 2.602 | (-11.856,-1.656) | **9.42E-03** |
| cg26312951 | 21 | 42797847 | *MX1* | 8 | 0.286 | 0.137 | (0.018,0.554) | **3.68E-02** | 12 | -4.582 | 1.261 | (-7.053,-2.111) | **2.79E-04** |
| cg22917487 | 3 | 39322103 | *CX3CR1* | 8 | 0.090 | 0.045 | (0.001,0.179) | **4.83E-02** | 7 | -3.522 | 5.577 | (-14.453,7.410) | 5.28E-01 |
| cg08122652 | 3 | 122281939 | *PARP9* | 8 | 0.259 | 0.141 | (-0.017,0.536) | 6.61E-02 | 8 | -4.517 | 1.558 | (-7.571,-1.463) | **3.75E-03** |
| cg22930808 | 3 | 122281881 | *PARP9* | 8 | 0.245 | 0.164 | (-0.076,0.567) | 1.35E-01 | 10 | -5.245 | 1.135 | (-7.470,-3.019) | **3.85E-06** |
| cg08818207 | 6 | 32820355 | *TAP1* | 8 | 0.092 | 0.071 | (-0.048,0.231) | 1.96E-01 | 2 | -10.287 | 5.519 | (-21.104,0.530) | 6.23E-02 |
| cg03038262 | 11 | 315262 | *IFITM1* | 8 | 0.099 | 0.081 | (-0.059,0.257) | 2.20E-01 | 4 | -9.610 | 3.955 | (-17.361,-1.859) | **1.51E-02** |
| cg22940798 | 6 | 32805554 | *TAP2* | 8 | 0.047 | 0.042 | (-0.035,0.128) | 2.62E-01 | 2 | -28.774 | 9.027 | (-46.466,-11.082) | **1.43E-03** |
| cg06981309 | 3 | 146260954 | *PLSCR1* | 8 | 0.107 | 0.097 | (-0.083,0.296) | 2.69E-01 | 4 | 1.502 | 2.983 | (-4.344,7.349) | 6.15E-01 |
| cg00096307 | 2 | 106761754 | *UXS1* | 8 | 0.036 | 0.033 | (-0.030,0.101) | 2.84E-01 | 2 | 3.455 | 11.853 | (-19.776,26.685) | 7.71E-01 |
| cg07839457 | 16 | 57023022 | *NLRC5* | 8 | 0.109 | 0.158 | (-0.200,0.418) | 4.88E-01 | 7 | -2.961 | 1.552 | (-6.004,0.081) | 5.64E-02 |
| cg26396492 | 20 | 19915762 | *RIN2* | 8 | 0.014 | 0.031 | (-0.047,0.075) | 6.50E-01 | 10 | -9.992 | 5.226 | (-20.234,0.250) | 5.59E-02 |
| cg08623256 | 5 | 3858275 |  | 8 | 0.009 | 0.029 | (-0.048,0.066) | 7.58E-01 | 1 | 18.246 | 15.315 | (-11.772,48.263) | 2.34E-01 |
| cg22385827 | 2 | 211036478 | *C2orf67* | 8 | 0.025 | 0.125 | (-0.219,0.269) | 8.41E-01 | 3 | 1.054 | 1.855 | (-2.582,4.689) | 5.70E-01 |

\*IV: instrumental variable
†p<0.05 are bolded

In step 2, we conducted the MR analysis based on cis-meQTLs of the candidate CpGs and their association with HIV severity in our sample. Seven CpGs showed significant MR estimates on HIV severity (**Table 5**). Of note, 3 significant CpGs in the MR analysis in step 1 were also significant in step 2.

Overall, 3 mediator CpGs discovered by the mediation analysis were validated as significant mediators by two-step epigenetic MR analysis (cg03753191,*EPSTI1;* cg06188083,*IFIT3;* cg26312951, *MX1*). Three CpGs without significant mediation effects in the mediation analysis were also found to be nonsignificant in the two-step MR analysis (cg26396492, *RIN2;* cg22385827,*C2orf67;* cg08623256).

## Discussion

Our findings provide evidence that cocaine use worsens HIV severity and increases mortality among HIV positive participants and that cocaine’s adverse effects are partially mediated by DNAm in the blood. We identified 12 CpGs that collectively accounted for a total of 47.2% of cocaine affecting HIV severity. Three of the 12 mediator CpGs were further validated by a two-step epigenetic MR approach, which provides supporting evidence that our mediation results were not affected by unmeasured confounders or reverse causation. The sensitivity analysis showed that our mediation analyses are relatively robust to slight violation of assumptions. These 12 mediator CpGs offer new insights into the mechanisms of how cocaine use may affect HIV outcomes by DNAm.

Methodological considerations are important for examining the mediation effect of DNAm. It is possible that our mediation analyses could be undermined by violation of model assumptions, reverse causation and unmeasured confounding. To address these concerns, we performed the sensitivity analysis and two-step epigenetic MR analysis to further evaluate the mediation effects of the 12 CpG sites. The results from the sensitivity test showed that the 12 mediator CpGs were robust when slight violation of the sequential ignorability assumption is present. The two-step epigenetic MR analysis confirmed 3 of 12 CpG sites as mediators of cocaine affecting HIV severity and were not affected by reverse causation and unmeasured confounding. Of note, the 8 SNPs used in the MR analysis showed marginal association with cocaine use, which limited their utility as instrumental variables and may explain why 9 CpG sites did not show significant mediation effects in two-step epigenetic MR analysis. In addition, in the mediation analysis, the HIV severity was measured after the blood collection for DNAm profiling to assure that the measurement of mediator precedes the measurement of outcome. Our study design intended to match the temporality of exposure, mediator and outcome and to avoid reverse causation. Of note, we observed discrepancy on the direction of cocaine use effect on DNA methylation between EWA scan and step 1 MR analysis. This may happen because EWA scan assessed association while MR evaluated the causal effect by removing reverse causality. This difference might also be due to different ways on adjusting for confounding factors in two models. Additionally, to assess whether cocaine use influenced cell type proportions as reflected by DNA methylation, we conducted a MR on cocaine affecting six cell type proportions using the same SNP instruments as used in step 1 MR. We found no significant MR estimates across six cell types (p>0.1) (**supplementary table S5**), suggesting that cocaine use does not directly affect cell type proportions in our sample. Overall, we took various measures to make sure our mediation results are valid and robust.

We observed that the sum of individual mediation proportion for 12 mediator CpGs exceeded 100%. An alternative approach is to test the joint mediation effect of all mediators [71]. We found that the 12 mediator CpGs jointly accounted for 47% of the total effect (effect size=0.329) of cocaine use on HIV severity. This finding indicates that these mediators may affect one another or that there is an interaction effect [71]. For example, several mediator CpG sites are near genes on the response to cytokine pathway *(PARP9, PLSCR1, CX3CR1, IFITM1, IFIT3, MX1* and *NLRC5)*. It is possible that these CpGs may share the common biological pathway on mediating the effect of cocaine on HIV severity.

These 12 CpGs are located in or near 11 biologically meaningful genes that were previously reported to be involved in inflammation, HIV-1 viral replication and other pathways that play critical roles in HIV progression. Specifically, cg06188083 on *IFIT3* mediated 28.8% of the variation, and *IFIT3* encodes an IFN-induced antiviral protein which acts as an inhibitor of viral processes and viral replication [76]. Another significant mediator CpG site, cg06188083, is located near interferon gene *IFITM1*. We previously reported the hypomethylation of cg07839457 due to HIV infection, which is located in the promoter region of *NLRC5* [33]. This CpG site was also a significant mediator between cocaine and HIV severity in this study. *NLRC5* plays an important role in the cytokine response and antiviral immunity through its inhibition of NF-kappa-B activation and negative regulation of type I interferon signaling pathways [77]. The converging evidence on cg07839457 *(NLRC5)* warrants further investigation of its role in HIV infection and progression. Another interesting CpG site, cg22917487 on *CX3CR1*, showed both strong association with persistent cocaine use and a significant mediation effect of cocaine affecting HIV severity. *CX3CR1* is involved in leukocyte adhesion and migration and was recently identified as an HIV-1 coreceptor [78]. Some studies also showed that genetic variants on *CX3CR1* were associated with HIV susceptibility and rapid HIV progression to AIDS [79]. cg25114611, located in the promoter region of *FKBP5*, is also biologically plausible, given the implication for chronic cocaine administration upregulating *FKBP5* expression in rats [80].

Cocaine use commonly cooccurred with the use of other substances, and this may confound cocaine’s effects on HIV severity and the mediation effects of CpGs between cocaine use and HIV outcomes. However, our results show that the association between cocaine use and HIV severity remained significant after accounting for smoking, alcohol, cannabis and opioid use. Additionally, our cocaine use EWA model adjusted for smoking as a covariate, and the selected candidate CpGs were not associated with alcohol, marijuana and opioid use (**Table S3**).

There are several strengths of this study. First, instead of selecting candidate mediator CpGs based on the literature or hypotheses, we applied an unbiased epigenome-wide screening to select CpGs associated with both cocaine use and HIV severity. Second, to limit self-reporting bias of cocaine use, we leveraged longitudinal data in defining persistent cocaine use and no cocaine use. We included only those participants who consistently reported cocaine use or no cocaine use across all 5 visits for the selection of candidate CpGs and the mediation analyses. Last, we used the average VACS index after blood collection so that DNAm measurements (mediator) preceded HIV severity (outcome) for the mediation analyses.

One limitation of the study is that our sample size for the mediation analyses is small. However, the strict definition of cocaine use helped reduce self-reporting bias and can potentially increase power by comparing extreme groups. In addition, we used a less stringent criterion when selecting candidate CpGs for mediation analysis due to limited sample size to achieve epigenome-wide significance. To our knowledge, there are no sufficiently sized independent cohorts for replication. Although this approach has also been adopted by previous studies [42, 81], using epigenome-wide significant CpG sites as candidate mediators may show stronger signals in the future study with a larger sample size. Additionally, other unmeasured confounding factors such as socioeconomic status may not be fully addressed in the mediation model. Lastly, our samples consisted of mostly male veterans, which may limit the generalizability of our findings.

## Conclusions

We validated previous reports that the use of cocaine worsened HIV severity and increased the risk of all-cause mortality among HIV positive participants. For the first time, this study found that several biologically meaningful DNAm sites mediated the adverse effect of cocaine use on HIV severity. These results merit future studies to further explore the biological mechanisms revealed by these DNAm sites on how cocaine affects HIV disease outcomes.

## Data Availability

Methylation data is available on GEO

https://www.ncbi.nlm.nih.gov/geo/query/acc.cgi?acc=GSE100264

---

**Figure S1**. Manhattan and quantile-quantile (QQ) plot of persistent cocaine use epigenome-wide association (EWA) (λ=1.034) and HIV severity EWA (λ=1.116)

**Figure S2**. The estimated ACME and their 95% confidence interval as a function of the sensitivity parameter ρ among 15 candidate CpGs

**List of abbreviations**
ACME: average causal mediation effect
ART: antiretroviral therapy
AUDIT-C: Alcohol Use Diagnosis Identification Test-consumption
CI: Confidence interval
DNA: Deoxyribonucleic acid
DNAm: DNA methylation
EWA: epigenome-wide association
FDR: False discovery rate
GWAS: Genome-wide association study
GO: Gene ontology
HIV: Human immunodeficiency virus
HM450K: Infinium Human Methylation 450K BeadChip
EPIC: Infinium Human Methylation EPIC BeadChip
HR: Hazard ratio
IL: interleukins
IVW: Inverse-variance weighted
LD: Linkage disequilibrium
meQTL: Methylation quantitative trait loci
MR: Mendelian randomization
NK: Natural killer
PC: Principal component
QC: Quality control
VACS: Veterans Aging Cohort Study
VACS-BC: Veteran Aging Cohort Study Biomarker Cohort
VACS index: Veterans Aging Cohort Study index

## Declarations

### Ethics approval and consent to participate

The study was approved by the committee of the Human Research Subject Protection at Yale University and the Institutional Research Board Committee of the Connecticut Veteran Healthcare System. All participants provided written consent.

### Availability of data and materials

Demographic and clinical variables and DNAm data for the VACS samples were submitted to the GEO dataset (GSE117861) and are available to the public. All codes for analysis are also available upon a request to the corresponding author.

### Competing interest

All authors declare that they have no conflict of interest.

### Funding

The work was supported by the National Institute on Drug Abuse (R03DA039745, R01DA038632, R01DA047063, R01DA047820).

### Authors’ contributions

CS was responsible for data analysis and manuscript preparation. ACJ provided DNA samples, clinical data, and contributed to manuscript preparation. XZ was responsible for the bioinformatics data processing. ZW contributed to mediation analysis and manuscript preparation. DH and EJ contributed to the analytical approach and the manuscript preparation. KX was responsible for the study design, study protocol, sample preparation, data analysis, interpretation of findings, and manuscript preparation.

## Acknowledgements

The authors appreciate the support of the Veteran Aging Study Cohort Biomarker Core and Yale Center of Genomic Analysis.

## Notes

### Competing Interest Statement

The authors have declared no competing interest.

### Summary of Updates

Revision according to reviewer's comments

## Reference

1. Daskalopoulou M, Rodger A, Phillips AN et al. Recreational drug use, polydrug use, and sexual behaviour in HIV-diagnosed men who have sex with men in the UK: results from the cross-sectional ASTRA study. Lancet HIV 1(1), e22–31 (2014).

2. Cofrancesco Jr J, Scherzer R, Tien PC et al. Illicit drug use and HIV treatment outcomes in a US cohort. AIDS (London, England) 22(3), 357 (2008).

3. Hatfield LA, Horvath KJ, Jacoby SM, Simon Rosser BR. Comparison of substance use and risky sexual behavior among a diverse sample of urban, HIV-positive men who have sex with men. J Addict Dis 28(3), 208–218 (2009).

4. Grabovac I, Meilinger M, Schalk H, Leichsenring B, Dorner TE. Prevalence and Associations of Illicit Drug and Polydrug Use in People Living with HIV in Vienna. Scientific Reports 8(1), 8046 (2018).

5. Wolitski RJ, Parsons JT, Gómez CA. Prevention with HIV-seropositive men who have sex with men: lessons from the Seropositive Urban Men’s Study (SUMS) and the Seropositive Urban Men’s Intervention Trial (SUMIT). J Acquir Immune Defic Syndr 37 Suppl 2 S101–109 (2004).

6. Purcell DW, Moss S, Remien RH, Woods WJ, Parsons JT. Illicit substance use, sexual risk, and HIV-positive gay and bisexual men: differences by serostatus of casual partners. Aids 19 Suppl 1 S37–47 (2005).

7. Key substance use and mental health indicators in the United States: Results from the 2018 National Survey on Drug Use and Health (HHS Publication No. PEP19-5068, NSDUH Series H-54). Rockville, MD: Center for Behavioral Health Statistics and Quality, Substance Abuse and Mental Health Services Administration. Retrieved from https://www.samhsa.gov/data/

8. Webber MP, Schoenbaum EE, Gourevitch MN, Buono D, Klein RS. A prospective study of HIV disease progression in female and male drug users. Aids 13(2), 257–262 (1999).

9. Vittinghoff E, Hessol NA, Bacchetti P, Fusaro RE, Holmberg SD, Buchbinder SP. Cofactors for HIV disease progression in a cohort of homosexual and bisexual men. Journal of acquired immune deficiency syndromes (1999) 27(3), 308–314 (2001).

10. Cook JA, Burke-Miller JK, Cohen MH et al. Crack cocaine, disease progression, and mortality in a multi-center cohort of HIV-1 positive women. AIDS (London, England) 22(11), 1355 (2008).

11. Baum MK, Rafie C, Lai S, Sales S, Page B, Campa A. Crack-cocaine use accelerates HIV disease progression in a cohort of HIV-positive drug users. JAIDS Journal of Acquired Immune Deficiency Syndromes 50(1), 93–99 (2009).

12. Sharpe TT, Lee LM, Nakashima AK, Elam-Evans LD, Fleming PL. Crack cocaine use and adherence to antiretroviral treatment among HIV-infected black women. J Community Health 29(2), 117–127 (2004).

13. Carrico AW, Johnson MO, Morin SF et al. Stimulant use is associated with immune activation and depleted tryptophan among HIV-positive persons on anti-retroviral therapy. Brain, behavior, and immunity 22(8), 1257–1262 (2008).

14. Dash S, Balasubramaniam M, Villalta F, Dash C, Pandhare J. Impact of cocaine abuse on HIV pathogenesis. Front Microbiol 6 1111–1111 (2015).

15. Rasbach DA, Desruisseau AJ, Kipp AM et al. Active cocaine use is associated with lack of HIV-1 virologic suppression independent of nonadherence to antiretroviral therapy: use of a rapid screening tool during routine clinic visits. AIDS care 25(1), 109–117 (2013).

16. Breitling LP, Yang R, Korn B, Burwinkel B, Brenner H. Tobacco-smoking-related differential DNA methylation: 27K discovery and replication. The American Journal of Human Genetics 88(4), 450–457 (2011).

17. Joubert BR, Håberg SE, Nilsen RM et al 450K epigenome-wide scan identifies differential DNA methylation in newborns related to maternal smoking during pregnancy. Environmental health perspectives 120(10), 1425–1431 (2012).

18. Lee KW, Pausova Z. Cigarette smoking and DNA methylation. Frontiers in genetics 4 132 (2013).

19. Tsaprouni LG, Yang T-P, Bell J et al. Cigarette smoking reduces DNA methylation levels at multiple genomic loci but the effect is partially reversible upon cessation. Epigenetics 9(10), 1382–1396 (2014).

20. Gao X, Zhang Y, Saum K-U, Schöttker B, Breitling LP, Brenner H. Tobacco smoking and smoking-related DNA methylation are associated with the development of frailty among older adults. Epigenetics 12(2), 149–156 (2017).

21. Zhang X, Hu Y, Aouizerat BE et al. Machine learning selected smoking-associated DNA methylation signatures that predict HIV prognosis and mortality. 10(1), 155 (2018).

22. Zhang R, Miao Q, Wang C et al. Genome-wide DNA methylation analysis in alcohol dependence. Addiction biology 18(2), 392–403 (2013).

23. Zhang H, Gelernter J. DNA methylation and alcohol use disorders: Progress and challenges. The American journal on addictions 26(5), 502–515 (2017).

24. Sharp GC, Arathimos R, Reese SE et al. Maternal alcohol consumption and offspring DNA methylation: findings from six general population-based birth cohorts. Epigenomics 10(1), 27–42 (2018).

25. Liu C, Marioni RE, Hedman ÅK et al. A DNA methylation biomarker of alcohol consumption. Molecular psychiatry 23(2), 422 (2018).

26. Joehanes R, Just AC, Marioni RE et al. Epigenetic Signatures of Cigarette Smoking. Circ Cardiovasc Genet 9(5), 436–447 (2016).

27. Feinberg AP, Koldobskiy MA, Göndör A. Epigenetic modulators, modifiers and mediators in cancer aetiology and progression. Nature Reviews Genetics 17 284 (2016).

28. Hao X, Luo H, Krawczyk M et al. DNA methylation markers for diagnosis and prognosis of common cancers. Proceedings of the National Academy of Sciences 114(28), 7414 (2017).

29. Michalak EM, Burr ML, Bannister AJ, Dawson MA. The roles of DNA, RNA and histone methylation in ageing and cancer. Nature Reviews Molecular Cell Biology doi:10.1038/s41580-019-0143-1 (2019).

30. Davegårdh C, García-Calzón S, Bacos K, Ling C. DNA methylation in the pathogenesis of type 2 diabetes in humans. Mol Metab 14 12–25 (2018).

31. Zhong J, Agha G, Baccarelli AA. The role of DNA methylation in cardiovascular risk and disease: methodological aspects, study design, and data analysis for epidemiological studies. Circulation research 118(1), 119–131 (2016).

32. Nakatochi M, Ichihara S, Yamamoto K et al. Epigenome-wide association of myocardial infarction with DNA methylation sites at loci related to cardiovascular disease. Clinical epigenetics 9(1), 54 (2017).

33. Zhang X, Justice AC, Hu Y et al. Epigenome-wide differential DNA methylation between HIV-infected and uninfected individuals. Epigenetics 11(10), 750–760 (2016).

34. Liu Y, Aryee MJ, Padyukov L et al. Epigenome-wide association data implicate DNA methylation as an intermediary of genetic risk in rheumatoid arthritis. Nature biotechnology 31(2), 142 (2013).

35. Bind M-A, Lepeule J, Zanobetti A et al. Air pollution and gene-specific methylation in the Normative Aging Study: association, effect modification, and mediation analysis. Epigenetics 9(3), 448–458 (2014).

36. Cao-Lei L, Dancause KN, Elgbeili G et al. DNA methylation mediates the impact of exposure to prenatal maternal stress on BMI and central adiposity in children at age 1334 years: Project Ice Storm. Epigenetics 10(8), 749–761 (2015).

37. Timms JA, Relton CL, Rankin J, Strathdee G, Mckay JA. DNA methylation as a potential mediator of environmental risks in the development of childhood acute lymphoblastic leukemia. Epigenomics 8(4), 519–536 (2016).

38. Tobi EW, Slieker RC, Luijk R et al. DNA methylation as a mediator of the association between prenatal adversity and risk factors for metabolic disease in adulthood. Science advances 4(1), eaao4364 (2018).

39. Barker ED, Walton E, Cecil CA. Annual Research Review: DNA methylation as a mediator in the association between risk exposure and child and adolescent psychopathology. Journal of Child Psychology and Psychiatry 59(4), 303–322 (2018).

40. Rutten BPF, Mill J. Epigenetic Mediation of Environmental Influences in Major Psychotic Disorders. Schizophrenia Bulletin 35(6), 1045–1056 (2009).

41. Ladd-Acosta C, Fallin MD. The role of epigenetics in genetic and environmental epidemiology. Epigenomics 8(2), 271–283 (2016).

42. Jordahl KM, Phipps AI, Randolph TW et al. Differential DNA methylation in blood as a mediator of the association between cigarette smoking and bladder cancer risk among postmenopausal women. Epigenetics doi:10.1080/15592294.2019.16311121-9 (2019).

43. Shirazi J, Shah S, Sagar D et al. Epigenetics, drugs of abuse, and the retroviral promoter. J Neuroimmune Pharmacol 8(5), 1181–1196 (2013).

44. Dhillon NK, Williams R, Peng F et al. Cocaine-mediated enhancement of virus replication in macrophages: implications for human immunodeficiency virus-associated dementia. J Neurovirol 13(6), 483–495 (2007).

45. Atluri VS, Pilakka-Kanthikeel S, Garcia G et al. Effect of Cocaine on HIV Infection and Inflammasome Gene Expression Profile in HIV Infected Macrophages. Sci Rep 6 27864 (2016).

46. Castro FOF, Silva JM, Dorneles GP et al. Distinct inflammatory profiles in HIV-infected individuals under ART using cannabis, cocaine or cannabis plus cocaine. AIDS doi:10.1097/QAD.0000000000002296 (2019).

47. Parikh N, Dampier W, Feng R et al. Cocaine alters cytokine profiles in HIV-1-infected African American individuals in the DrexelMed HIV/AIDS genetic analysis cohort. JAcquir Immune Defic Syndr 66(3), 256–264 (2014).

48. Robison AJ, Nestler EJ. Transcriptional and epigenetic mechanisms of addiction. Nat Rev Neurosci 12(11), 623–637 (2011).

49. Anier K, Malinovskaja K, Aonurm-Helm A, Zharkovsky A, Kalda A. DNA methylation regulates cocaine-induced behavioral sensitization in mice. Neuropsychopharmacology 35(12), 2450–2461 (2010).

50. Camilo C, Maschietto M, Vieira HC et al. Genome-wide DNA methylation profile in the peripheral blood of cocaine and crack dependents. BrazJ Psychiatry 41(6), 485–493 (2019).

51. Imai K, Keele L, Tingley D. A general approach to causal mediation analysis. Psychological methods 15(4), 309 (2010).

52. Relton CL, Davey Smith G. Two-step epigenetic Mendelian randomization: a strategy for establishing the causal role of epigenetic processes in pathways to disease. IntJ Epidemiol 41(1), 161–176 (2012).

53. Justice AC, Dombrowski E, Conigliaro J et al. Veterans aging cohort study (VACS): overview and description. 44(8 Suppl 2), S13 (2006).

54. Veterans Aging Cohort Study. VACS Biomarker Cohort Description. (2016).

55. Tate JP, Justice AC, Hughes MD et al. An internationally generalizable risk index for mortality after one year of antiretroviral therapy. AIDS (London, England) 27(4), 563–572 (2013).

56. Justice AC, Modur SP, Tate JP et al. Predictive accuracy of the Veterans Aging Cohort Study index for mortality with HIV infection: a North American cross cohort analysis. Journal of acquired immune deficiency syndromes (1999) 62(2), 149–163 (2013).

57. Bebu I, Tate J, Rimland D et al. The VACS Index Predicts Mortality in a Young, Healthy HIV Population Starting Highly Active Antiretroviral Therapy. J. Acquir. Immune. Defic. Syndr. 65(2), 226–230 (2014).

58. Brown ST, Tate JP, Kyriakides TC et al. The VACS index accurately predicts mortality and treatment response among multi-drug resistant HIV infected patients participating in the options in management with antiretrovirals (OPTIMA) study. PLoS One 9(3), e92606 (2014).

59. Justice AC, Modur S, Tate JP et al. Predictive accuracy of the Veterans Aging Cohort Study (VACS) index for mortality with HIV infection: a north American cross cohort analysis. Journal of acquired immune deficiency syndromes (1999) 62(2), 149 (2013).

60. Lehne B, Drong AW, Loh M et al. A coherent approach for analysis of the lllumina HumanMethylation450 BeadChip improves data quality and performance in epigenome-wide association studies. 16(1), 37 (2015).

61. Aryee MJ, Jaffe AE, Corrada-Bravo H et al. Minfi: a flexible and comprehensive Bioconductor package for the analysis of Infinium DNA methylation microarrays. 30(10), 1363–1369 (2014).

62. Houseman EA, Accomando WP, Koestler DC et al. DNA methylation arrays as surrogate measures of cell mixture distribution. BMC Bioinformatics 13(1), 86 (2012).

63. Johnson WE, Li C, Rabinovic A. Adjusting batch effects in microarray expression data using empirical Bayes methods. Biostatistics 8(1), 118–127 (2007).

64. Howie BN, Donnelly P, Marchini J. A flexible and accurate genotype imputation method for the next generation of genome-wide association studies. PLoS Genet 5(6), e1000529 (2009).

65. Purcell S, Neale B, Todd-Brown K et al. PUNK: a tool set for whole-genome association and population-based linkage analyses. The American journal of human genetics 81(3), 559–575 (2007).

66. Kassambara A, Kosinski M, Biecek P. survminer: Drawing Survival Curves using’ggplot2’. R package version 0.3 1 (2017).

67. Therneau T. A Package for Survival Analysis in S. version 2.38. (2015).

68. Zhang X, Hu Y, Justice AC et al. DNA methylation signatures of illicit drug injection and hepatitis C are associated with HIV frailty. Nature Communications 8(1), 2243 (2017).

69. Jaffe A. Flowsorted. blood. 450k: illumina humanmethylation data on sorted blood cell populations. R Package Version 1(0), (2015).

70. Tingley D, Yamamoto T, Hirose K, Keele L, Imai K. Mediation: R package for causal mediation analysis. (2014).

71. Vanderweele T, Vansteelandt S. Mediation analysis with multiple mediators. Epidemiologic methods 2(1), 95–115 (2014).

72. Yavorska OO, Burgess S. MendelianRandomization: an R package for performing Mendelian randomization analyses using summarized data. International Journal of Epidemiology 46(6), 1734–1739 (2017).

73. Cabana-Domínguez J, Shivalikanjli A, Fernàndez-Castillo N, Cormand B. Genome-wide association meta-analysis of cocaine dependence: Shared genetics with comorbid conditions. Progress in Neuro-Psychopharmacology and Biological Psychiatry 94 109667 (2019).

74. Myers TA, Chanock SJ, Machiela MJ. LDlinkR: An R Package for Rapidly Calculating Linkage Disequilibrium Statistics in Diverse Populations. Frontiers in Genetics 11157 (2020).

75. Auton A, Abecasis GR, Altshuler DM et al. A global reference for human genetic variation. Nature 526(7571), 68–74 (2015).

76. Schmeisser H, Mejido J, Balinsky CA et al. Identification of alpha interferon-induced genes associated with antiviral activity in Daudi cells and characterization of IFIT3 as a novel antiviral gene. J Virol 84(20), 10671–10680 (2010).

77. Cui J, Zhu L, Xia X et al. NLRC5 negatively regulates the NF-kappaB and type I interferon signaling pathways. Cell 141(3), 483–496 (2010).

78. Garin A, Tarantino N, Faure S et al. Two novel fully functional isoforms of CX3CR1 are potent HIV coreceptors. J Immunol 171(10), 5305–5312 (2003).

79. Faure S, Meyer L, Costagliola D et al. Rapid progression to AIDS in HIV+ individuals with a structural variant of the chemokine receptor CX3CR1. Science 287(5461), 2274–2277 (2000).

80. Connelly KL, Unterwald EM. Chronic cocaine administration upregulates FKBP5 in the extended amygdala of male and female rats. Drug Alcohol Depend 199 101–105 (2019).

81. Chu SH, Loucks EB, Kelsey KT et al. Sex-specific epigenetic mediators between early life social disadvantage and adulthood BMI. Epigenomics 10(6), 707–722 (2018).

